# Knowledge about COVID-19 and patients’ beliefs about and use of herbal products during the COVID-19 pandemic: a cross-sectional study in Saudi Arabia

**DOI:** 10.1101/2020.06.23.20138107

**Authors:** Hamad S. Alyami, Mohamed A. A. Orabi, Fahad M. Aldhabbah, Hamad N. Alturki, Wejdan I. Aburas, Abdulrahman I. Alfayez, Abdullah S. Alharbi, Rawan A. Almasuood, Njood A. Alsuhaibani

**Affiliations:** Department of Pharmaceutics, College of Pharmacy, Najran University, Najran, Saudi Arabia; Department of Pharmacognosy, College of Pharmacy, Najran University, Najran, Saudi Arabia; Department of Pharmacognosy, Faculty of Pharmacy, Al-Azhar University, Assiut-branch, Assiut, Egypt; College of Pharmacy, Qassim University, Al-Qassim, Saudi Arabia; College of Pharmacy, University of Hail, Hail, Saudi Arabia

**Keywords:** COVID-19, Food supplements, Herbal products, Saudi Arabia

## Abstract

The lack of treatment options for COVID-19 has raised many concerns among populations worldwide, which has led to many attempts to find alternative options to prevent the transmission of the disease or to alleviate the progression of the infection, including focusing more on preventive measures (to prevent transmission to other individuals) and the use of natural products and herbal extracts to increase immunity and decrease the probability of getting infected. This study explored the knowledge of the population of the Kingdom of Saudi Arabia (KSA) about COVID-19 preventive measures and their belief about the consumption of herbal products for the prevention of COVID-19 infection. A total of 5,258 individuals participated in this study. Participants’ knowledge about the appropriate COVID-19 preventive measures in terms of handwashing procedures, self-quarantine and social distancing was moderate, with a mean score of 5.5 (SD: 1.7) out of 10 (representing around 55.0% of the total score). About 22.1% (n= 1,161) of the participants reported that they have used herbal products or nutritional supplements during the pandemic period, to protect themselves from the disease. Social media and the Internet (39.4%, n= 372) were the main motivators for the participants to try herbal products. Vitamin C was the most commonly used food supplement to increase immunity and reduces the chance of contracting COVID-19. Our study demonstrated that the general population in Saudi Arabia has a moderate level of knowledge about COVID-19 transmission and preventive measures. A considerable proportion of the population reported the use of herbal products or food supplements in order to protect themselves from the disease. Policymakers are suggested to provide further educational campaigns that increase population knowledge about the disease transmission routes and preventive measures. In addition, the use of herbal products should be evidence-based to ensure patient safety.

## 1. Introduction

On December 31, 2019, several patients were admitted to the local hospitals of Wuhan, China, with symptoms of pneumonia linked to a common infection in the wholesale market in Wuhan. COVID-19 symptoms include fever, cough, fatigue, diarrhoea, and headache. In addition to that, the transmission of the virus is noticed to be fast in direct or indirect pathways, such as respiratory droplets, direct contact with an infected person, and on surfaces and objects, which has led to more than 5 million cases of COVID-19 worldwide and a death rate that has exceeded 300,000 deaths (Rothan and Byrareddy 2020; Shereen et al. 2020; World Health Organization 2020b; Liu et al. 2020; World Health Organization 2020a, 2020c). Researchers have compared the current COVID-19 pandemic to the Spanish flu that occurred in 1918, which resulted in more than 50 million deaths around the world. In addition to that, the rapid transmission rate and the lack of vaccinations or any treatment option for the COVID-19 outbreak provoked the World Health Organisation (WHO) and the International Health Regulation (IHR) emergency committee to announce the case as a Public Health Emergency of International Concern (PHEIC) on January 30, 2020 (Centers for Disease Control and Prevention 2019; Martini et al. 2019; Barry, Al Amri, and Memish 2020). Alarmingly, the investigation reports indicated a new strain of coronavirus with unknown aetiology that had spread globally, which forced the World Health Organization to declare it a pandemic in March 2020. It was named coronavirus disease 2019 (COVID-19) (Bogoch et al. 2020; World Health Organization 2020f; Zhao et al. 2020; Lu, Stratton, and Tang 2020). In March 2020, Saudi Arabia reported the first case of COVID-19, and the disease started to spread increasingly throughout the whole country. As a consequence, in order to prevent the rapid transmission of the infection, the Saudi government implemented multiple measures, including restrictions in the educational sectors and broad restrictions in the transportation sector (nationally and internationally). Subsequently, a general curfew was placed on the residents and citizens of Saudi Arabia in an attempt to curtail the spread of the virus (Saudi Ministry of Health 2020; Saudi Center for Disease Prevention and Control 2020). However, the transmission of the disease dramatically increased and reached more than 145,000 confirmed COVID-19 cases in the kingdom by the 18^th^ of June (Saudi Ministry of Health 2020).

The lack of treatment options for COVID-19 has raised many concerns among populations worldwide, which has led to many attempts to find alternative options to prevent the transmission of the disease or to alleviate the progression of the infection, including focusing more on preventive measures (to prevent transmission to other individuals) and the use of natural products and herbal extracts to increase immunity and decrease the probability of getting infected (Austin, Martin, and Gregory 2007; Lake 2020; World Health Organization 2020a, 2020c; Salathe et al. 2020).

Historically, the use of natural products was known to be the only treatment method against many infections and diseases. Also, natural products were used to enhance the immune system and are still widely used nowadays, mainly without the need for a prescription [19, 20]. Regardless of the benefits of using herbal products, there is a significant disadvantage, namely the herbal-drug interaction. Posadzki et al. (2013) reported the possible interactions between herbal medicinal products and drugs, which they classified as minor, moderate, and severe. Their study showed that, in some cases, the combination of herbal medicine and drugs resulted in life-threatening conditions (Posadzki, Watson, and Ernst 2013). Unfortunately, a recent report showed that about 80% of developed countries’ populations still rely on herbal medicines without any control (Pathak and Das 2013). Regarding COVID-19, many studies have been conducted to investigate the ability of already-known antiviral herbal products to treat it. In addition, there has been a dramatic increase in the use of this kind of medicine among individuals who are afraid of contracting COVID-19, without taking into consideration the possible harmful effects of some products (Ekor 2014). The objective of this study is to explore the knowledge of the population of the Kingdom of Saudi Arabia (KSA) about COVID-19 preventive measures and their belief about the consumption of herbal products for the prevention of COVID-19 infection.

## 2. Methods

### 2.1. Study design and study population

A cross-sectional study using an online survey was conducted in Saudi Arabia between May 23^rd^ and June 06^th^, 2020. The survey instrument was developed based on extensive literature reviews on the COVID-19 pandemic, including its transmission and preventive measures and the use of traditional medicine (World Health Organization 2020e; Centers for Disease Control and Prevention 2020; Luo et al. 2020). The questionnaire tool was reviewed by three researchers to evaluate the appropriateness, relevancy, clarity, and adequacy of the questions. The survey instrument included thirty-three questions, including socio-demographic characteristics, history of chronic diseases, respondents’ adherence to the Ministry of Health (MOH) instructions on COVID-19 pandemic preventive measures and social distancing, and the use of herbal products and food supplements. The survey was divided into five major sections. The first section had eight questions on the respondents’ sociodemographic characteristics. The second section included two questions about having a history of chronic diseases and whether the participant is currently/previously infected with COVID-19 (a yes-no question). The third section contained five questions on participants’ knowledge about COVID-19 preventive measures (specifically handwashing and social distancing). Each question was given a score of two for each correct answer (a total of 10 for the whole knowledge section). The total possible score could range between 0 – 10, and can be interpreted based on the mid-point of the highest possible score of the scale (equal to 5). The higher the score, the better the knowledge about COVID-19 preventive measures.

The knowledge section was followed by a question asking the participants whether they consider themselves committed to Saudi Ministry of Health recommendations regarding COVID-19 prevention and transmission. The fourth section (17 questions) asked them about their beliefs about herbal products and food supplements in terms of: 1) usage, 2) the source of information about them and the source of supply, and 3) belief in the protective effects of herbal products and food supplements as effective protection against COVID-19.

### 2.2. Sampling strategy

A convenient sample of eligible participants was invited to participate in the study through social media (Facebook, Twitter, Snapchat, Instagram and WhatsApp). All participants voluntarily participated in the study and were thus considered exempt from written informed consent. The study aims and objectives were clearly explained at the beginning of the survey.

The inclusion criteria were: a) participants aged 18 years and above and currently living in Saudi Arabia. Participants were excluded if they were: a) below 18 years of age; and b) unable to understand the Arabic language.

### 2.3. Sample size

Using a confidence interval of 95%, a standard deviation of 0.5, and a margin of error of 5%, the required sample size was 385 participants.

### 2.4. Statistical analysis

Data were analysed using SPSS software, version 25 (IBM Corp, Armonk, NY, USA). Continuous variables were reported as mean (± standard deviation [SD]), while categorical variables were reported as frequencies and percentages. The Kolmogorov–Smirnov and Shapiro Wilk tests were used to check the normality of the data. Participants’ knowledge scores were interpreted as a continuous scale based on the scale midpoint, where scores above the midpoint represented better knowledge about COVID-19 preventive measures. Student t-test and one-way ANOVA test were used to compare the mean knowledge scores between different demographic groups respectively. Tukey’s HSD post-hoc test was conducted to identify the source of significant variation within each group. Additionally, significant predictors of better knowledge were determined using multiple linear regression analysis. A confidence interval of 95% (p<0.05) was applied to represent the statistical significance of the results, and the level of significance was predetermined as 5%.

### 2.5. Ethical considerations

This study was approved by the general directorate of health affairs – Al-Qassem Region, Ministry of Health, Saudi Arabia, registration No H-04-Q-001. As participation in the study was voluntary, the research ethics committee approved the consent waiver.

## 3. Results

### 3.1. Characteristics of study participants

A total of 5,258 individuals participated in this study. More than half (57.1%, n= 3,000) were females. Around 41.0% (n= 2,155) of the participants were aged 25 to 39 years. The majority were married (60.3%, n= 3,169), Saudi (96.5%, n= 5,076), and living in central provinces (58.0%, n= 3,048) of the Kingdom. More than half of them (60.0%, n= 3,155) were bachelor degree holders, 45.4% (n= 2,387) were employed, and 34.3% (n= 1,803) have a family member working in the healthcare provision sector. Around 14.5% (n= 760) of them reported that they have a history of chronic disease and 0.6% (n= 30) reported that they had been or were currently infected with COVID-19. When the participants were asked whether they consider themselves committed to the Ministry of Health instructions regarding the COVID-19 pandemic, 43.0% (n= 2,262) reported that they are completely committed and 32.0% (n= 1,685) reported that they are committed most of the time except in emergency cases, refer to (**Table 1)**.

**Table 1.** Demographic characteristics of the study participants (n = 5,258).

| Characteristics | Frequency (%) |
| --- | --- |
| **Gender** |  |
| Female | 3,000 (57.1) |
| **Age categories (year)** |  |
| 18 - 24 | 1,161 (22.1) |
| 25 - 39 | 2,155 (41.0) |
| 40 - 59 | 1,801 (34.3) |
| 60 and above | 141 (2.7) |
| **Marital status** |  |
| Single | 1,885 (35.9) |
| Married | 3,169 (60.3) |
| Divorced | 135 (2.6) |
| Widowed | 69 (1.3) |
| **Nationality** |  |
| Saudi | 5,076 (96.5) |
| Non-Saudi | 182 (3.5) |
| **Residence area** |  |
| Central province | 3,048 (58.0) |
| Eastern province | 625 (11.9) |
| Western province | 607 (11.5) |
| Northern province | 504 (9.6) |
| Southern province | 474 (9.0) |
| **Education level** |  |
| Illiterate | 10 (0.2) |
| Lower than secondary level | 233 (4.4) |
| Secondary education or Diploma | 1,365 (26.0) |
| Bachelor degree | 3,155 (60.0) |
| Higher education | 495 (9.4) |
| **Employment status** |  |
| Student | 976 (18.6) |
| Healthcare-related career | 306 (5.8) |
| Employee | 2,387 (45.4) |
| Unemployed (retired or housewife) | 1,589 (30.1) |
| **Chronic disease history** |  |
| Yes | 760 (14.5) |
| **Where infected/currently infected with COVID-19?** |  |
| Yes | 30 (0.6) |
| **Level of commitment to MOH instructions regarding COVID-19 pandemic** |  |
| Full commitment | 2,262 (43.0) |
| Committed except in emergency cases | 1,685 (32.0) |
| Committed to some extent | 1,262 (24.0) |
| Not committed at all | 49 (0.9) |

### 3.2 Participants’ knowledge about COVID-19 preventive measures

Participants’ knowledge about the appropriate COVID-19 preventive measures in terms of handwashing procedures, self-quarantine and social distancing was moderate, with a mean score of 5.5 (SD: 1.7) out of 10 (representing around 55.0% of the total score).

The majority of the participants (90.1%, n= 4,736) agreed that washing hands with soap and water reduces the spread of coronavirus. Only 24.3% (n= 1,277) of the participants knew that handwashing should last at least 20 seconds, and 21.6% (n= 1,136) knew that there are five main steps in washing one’s hands correctly. The majority of the participants (95.9%, n= 5,044) reported that their commitment to staying at home and away from gatherings contributes to reducing COVID-19 transmission. However, only 40.1% (n= 2,107) of them were aware of the appropriate physical social distance (which is a minimum of one metre) required to prevent COVID-19 transmission.

#### 3.2.1. Participants’ demographics and their knowledge about COVID-19 preventive measures

**(Table 2)** presents the mean score for participants’ knowledge about COVID-19 preventive measures. Participants’ scores differed significantly by gender, age, education level, employment status and their level of commitment to MOH instructions regarding the COVID-19 pandemic. Females, individuals aged below 39 years, individuals with secondary school education or higher, university students, individuals who have reported that they are committed to MOH instructions regarding the COVID-19 pandemic, and users of herbal products and food supplements had a higher knowledge score compared to others (p< 0.05). The Tukey HSD test confirmed that those aged 60 years and above, those with higher education, employees, and those who reported that they are not committed at all were the main sources of significant variation between groups.

**Table 2.** COVID-19 mean knowledge score stratified by demographic characteristics.

| Demographic variable | Mean | SD | P-value |
| --- | --- | --- | --- |
| Gender |  |  |  |
| Female | 5.7 | 1.7 | 0.000*** |
| Male | 5.4 | 1.6 |  |
| Age category (Years) |  |  |  |
| 18 - 24 | 5.7 | 1.7 | 0.011* |
| 25 - 39 | 5.6 | 1.7 |  |
| 40 - 59 | 5.5 | 1.7 |  |
| 60 and above | 5.2 | 1.7 |  |
| Marital status |  |  |  |
| Single | 5.6 | 1.7 | 0.342 |
| Married | 5.5 | 6.0 |  |
| Divorced | 5.3 | 1.7 |  |
| Widowed | 5.5 | 1.9 |  |
| Nationality |  |  |  |
| Saudi | 5.5 | 1.7 | 0.402 |
| Non-Saudi | 5.7 | 1.7 |  |
| Residence area |  |  |  |
| Central province | 5.5 | 1.6 | 0.778 |
| Eastern province | 5.5 | 1.7 |  |
| Southern province | 5.5 | 1.6 |  |
| Western province | 5.6 | 1.7 |  |
| Northern province | 5.6 | 1.7 |  |
| Education level |  |  |  |
| Illiterate | 4.6 | 1.3 | 0.014* |
| Under secondary level | 5.6 | 1.7 |  |
| Secondary/Diploma | 5.6 | 1.6 |  |
| Bachelor | 5.6 | 1.7 |  |
| Higher education (Master/PhD) | 5.4 | 1.6 |  |
| Employment status |  |  |  |
| Student | 5.7 | 1.7 | 0.023* |
| Health-related career employee | 5.5 | 1.6 |  |
| Employee | 5.5 | 1.7 |  |
| Unemployed (retired or housewife) | 5.5 | 1.6 |  |
| Chronic disease history |  |  |  |
| Yes | 5.6 | 1.7 | 0.134 |
| No | 5.5 | 1.6 |  |
| Where infected/currently infected with COVID-19? |  |  |  |
| Yes | 5.3 | 2.0 | 0.401 |
| No | 5.6 | 1.7 |  |
| Have family member working in the healthcare provision sector? |  |  |  |
| Yes | 5.5 | 1.6 | 0.081 |
| No | 5.6 | 1.7 |  |
| Level of commitment to MOH instructions regarding COVID-19 pandemic |  |  |  |
| Full commitment | 5.6 | 1.6 | 0.000*** |
| Committed except in emergency cases | 5.7 | 1.6 |  |
| Committed to some extent | 5.4 | 1.7 |  |
| Not committed at all | 4.2 | 2.3 |  |
| Do you use or have you ever used herbal products or nutritional supplements during the COVID-19 pandemic period with the intention of protecting you from the disease? |  |  |  |
| Yes | 5.6 | 1.7 | 0.039* |
| No | 5.5 | 1.6 |  |

\*P< 0.05, \*\*\* P< 0.000.

Multiple linear regression analysis showed that being female, having secondary school education or lower, or higher education (PhD or a master’s degree) were important predictors of having a higher knowledge score about COVID-19 preventive measures (p< 0.05). On the other hand, being unemployed was an important predictor of having a relatively low knowledge score (p< 0.05), refer to (**Table 3)**.

**Table 3.** Multiple linear regression analysis predicting participants’ knowledge score.

| Variable | Model 1<sup>a</sup> |  |  | Model 2<sup>b</sup> |  |  |
| --- | --- | --- | --- | --- | --- | --- |
|  | B | SE | β | B | SE | β |
| **Demographic data** |  |  |  |  |  |  |
| **Age (years)** |  |  |  |  |  |  |
| 25 - 39 | 0.005 | 0.092 | 0.001 | 0.007 | 0.092 | 0.002 |
| 40 - 59 | -0.068 | 0.107 | -0.019 | -0.056 | 0.108 | -0.016 |
| 60 and above | -0.247 | 0.182 | -0.024 | -0.214 | 0.185 | -0.020 |
| **Male** | -0.373 | 0.052 | -0.111*** | -0.374 | 0.052 | -0.111*** |
| **Marital status** |  |  |  |  |  |  |
| Married | 0.069 | 0.073 | 0.021 | 0.069 | 0.073 | 0.021 |
| Divorced | -0.279 | 0.162 | -0.027 | -0.274 | 0.162 | -0.026 |
| Widowed | 0.062 | 0.222 | 0.004 | 0.067 | 0.222 | 0.005 |
| **Education level** |  |  |  |  |  |  |
| Under secondary level | 1.092 | 0.533 | 0.142* | 1.075 | 0.533 | 0.140* |
| Secondary/Diploma | 1.074 | 0.524 | 0.293* | 1.062 | 0.524 | 0.290* |
| Bachelor | 0.993 | 0.524 | 0.283 | 0.979 | 0.524 | 0.279 |
| Higher education (Master/PhD) | 0.209 | 0.078 | 0.037** | 0.131 | 0.079 | 0.023 |
| **Employment status** |  |  |  |  |  |  |
| Health-related career employee | -0.109 | 0.133 | -0.015 | -0.108 | 0.133 | -0.014 |
| Employee | -0.134 | 0.100 | -0.040 | -0.135 | 0.100 | -0.040 |
| Unemployed (retired or housewife) | -0.235 | 0.097 | -0.066* | -0.233 | 0.097 | -0.065* |
| **Having chronic disease history** |  |  |  |  |  |  |
| **Constant** | 4.838 | 0.525 |  | 4.855 | 0.526 |  |
| **Adjusted R<sup>2</sup>** |  |  | 0.012 |  |  | 0.012 |
| **P-value** |  |  | 0.000 |  |  | 0.000 |

a: includes age, gender, marital status, educational level, and employment status.
b: includes age, gender, marital status, educational level, employment status, and having chronic disease history.
B: the average change in the dependent variable associated with a 1 unit change in the independent variable, statistically controlling for the other independent variables;
SE: it is the standard deviation of its sampling distribution or an estimate of that standard deviation; β: a statistical measure that compares the strength of the effect of each individual independent variable to the dependent variable

### 3.3 Beliefs about the use of herbal products and/or food supplements as a protective measure

Around 14.9% (n= 783) of the participants reported that they have used herbal products or nutritional supplements during the pandemic period, to protect themselves from the disease. In addition, 7.2% (n= 378) reported that they had used these products for a while and then stopped.

Social media and the Internet (39.4%, n= 372) were the main motivators for the participants to try herbal products, and 37.5% (n= 354) of the participants reported that they obtained the herbal products from their own home. When they were asked about the types of dietary supplements that they have used or were currently using, 94.4% (n= 521) reported taking vitamin C. In terms of participants’ beliefs about the use of herbal products and/or food supplements as a protective measure, 34.4% of the participants believe that eating garlic helps to increase immunity and reduces the chance of contracting COVID-19. This belief in herbal products was followed by vitamin C, which 29.3% of the participants’ believe has a role in treating or reducing the chances of contracting COVID-19. For further details about beliefs regarding the use of herbal products and/or food supplements as a protective measure, refer to (**Table 4)**.

**Table 4.** Belief about the use of the herbal products and/or food supplements as protective measure.

| Variable | Frequency (%) |  |  |  |
| --- | --- | --- | --- | --- |
| **Do you use herbal products or supplements during the period of the pandemic in order to protect you from the disease?** (n= 5,245) |  |  |  |  |
| Yes | 783 (14.9) |  |  |  |
| No | 4,084 (77.9) |  |  |  |
| I used it for a while and then stopped it | 378 (7.2) |  |  |  |
| **If yes, who suggested you take a nutritional / herbal supplement?** (n= 944) |  |  |  |  |
| Dietician / physician / pharmacist / nurse / health practitioner | 223 (23.6) |  |  |  |
| Social media and other websites | 372 (39.4) |  |  |  |
| Recommendation from friend or relative | 349 (37.0) |  |  |  |
| **If yes, where did you get it?** |  |  |  |  |
| Herbal product shop | 293 (31.0) |  |  |  |
| Pharmacy | 226 (23.9) |  |  |  |
| Home | 354 (37.5) |  |  |  |
| Internet | 71 (0.07) |  |  |  |
| **What type of dietary supplement did you use before/use now during the pandemic?** (You can choose more than one answer) (n= 552) |  |  |  |  |
| Vitamin C | 521 (94.4) |  |  |  |
| Royal Jelly | 31 (5.6) |  |  |  |
| **Beliefs about the use of the herbal products and/or food supplements as protective measure** |  |  |  |  |
|  | **Yes** | **No** | **Maybe** | **I don't know** |
| Do you think that drinking turmeric tea helps increase immunity and reduce the chance of developing COVID-19? | 13.3% | 19.6% | 25.1% | 42.0% |
| Do you think ginger tea helps to increase the immunity and reduce the chance of developing COVID-19? | 19.5% | 17.5% | 33.0% | 30.0% |
| Do you think that eating garlic helps to increase the immunity and reduce the chance of developing COVID-19? | 34.4% | 14.8% | 32.6% | 18.3% |
| Do you think that eating onions (or onion peel) helps to increase the immunity and reduce the chance of developing COVID-19? | 20.6% | 20.1% | 28.8% | 30.5% |
| Do you think that eating fish oil known as omega-3 helps to increase the immunity and reduce the chance of developing COVID-19? | 9.5% | 19.8% | 25.6% | 45.1% |
| Do you think that taking ginseng extract tablets help to increase the immunity and protect us from corona infection? | 3.4% | 20.9% | 16.5% | 59.2% |
| Do you think that consumption of vitamin C found in citrus has a role in treating or reducing the chances of developing COVID-19? | 29.3% | 13.7% | 33.4% | 23.5% |
| Do you think that vinegar plays a role in treating or protecting against the COVID-19? | 5.9% | 29.3% | 18.7% | 46.1% |
| Do you think salted water plays a role in treating or protecting against the COVID-19? | 10.0% | 33.4% | 21.4% | 35.2% |
| Do you think that applying Sesame oil on the body protect from COVID-19? | 1.2% | 51.6% | 6.9% | 40.3% |
| Do you think that consuming of nutritional and herbal supplements prevents the spread of COVID-19 more than | 15.7% | 23.6% | 32.2% | 28.5% |
| social distance? |  |  |  |  |
| Do you think that vitamins and herbal supplements<br>treat/reduce the incidence of COVID-19? | 4.4% | 57.3% | 14.0% | 24.3% |

## 3. Discussion

This study explored beliefs about and the use of herbal products and food supplements among the general population of the Kingdom of Saudi Arabia. In addition, this study explored the population’s knowledge about COVID-19 preventive measures, specifically handwashing techniques and social distancing. Besides this, the level of knowledge about COVID-19 was assessed between users and non-users of herbal products and food supplements.

The findings of our study highlight the importance of educating the general population about COVID-19 transmission and the preventive measures that play a vital role in their protection from getting infected with the disease and which dramatically reduce the spread of the disease.

The explosive spread of COVID-19 has been a matter of international concern, including in the Middle Eastern countries. The SARS-CoV-2 high infectivity rate has been established regardless of the appearance of symptoms, which may extend to 14 days (Algaissi et al. 2020; Singhal 2020). Current evidence indicates that the causative virus for COVID-19 is transmitted through respiratory droplets or when contaminated hands touch the mucosa of the mouth, nose, or eyes; the virus can also be transferred from one surface to another by contaminated hands, which facilitates indirect contact transmission. These facts, together with the lack of effective vaccines or treatments, have pushed the health authorities to consider many preventive measures to stem the spread of the pandemic (Nemati, Ebrahimi, and Nemati 2020; Sen-Crowe, McKenney, and Elkbuli 2020). In the Kingdom of Saudi Arabia, the Ministry of Health, based on its previous experience from the Middle East Respiratory Syndrome (MERS) epidemic in 2012, has immediately established multiple campaigns, launched mobile health applications and call centres, and provided the public with information about COVID-19, as well as providing answers to related queries to enhance public awareness and engage the public in active learning and searching for information about related precautionary measures (Algaissi et al. 2020). These approaches have provided residents with information related to washing hands with soap in the correct way for the required amount of time, the benefits of staying at home, and related precautionary measures to prevent the spread of the COVID-19 virus (Aldarhami et al. 2020). The majority of the participants (75.0%, n= 3,947) reported that they are committed (either all the time or except in some emergency circumstances) to the recommendations of the Saudi Ministry of Health regarding the COVID-19 pandemic, which could reflect the effectiveness of the timely organisation of plans (even before the pandemic reached KSA) by the Saudi MOH to raise awareness among the population about preventing the spread of COVID-19. This is probably due to the previous experience of the Saudi people in response to the Middle East Respiratory Syndrome (MERS) epidemic in 2012 (Aldarhami et al. 2020). Nevertheless, the level of knowledge among the population about COVID-19 preventive measures was moderate (55.0%; 5.5 out of 10). This confirms the result of a recent study that was conducted among three Arabic countries (Saudi Arabia, Jordan and Kuwait), which reported that the overall population knowledge about COVID-19 was moderate (66.1%). Furthermore, and confirming the findings of our study, this study highlighted that the population knowledge is mainly defective as regards disease transmission routes (Naser et al. 2020). Less than 25.0% of the participants knew the proper procedure of handwashing in terms of the time needed and the proper procedure itself. In addition, only 40.1% (n= 2,107) of them were aware of the appropriate physical social distance needed to protect from COVID-19 transmission. The World Health Organization has emphasised the importance of taking actions that optimise protection and control of disease transmission among the community (World Health Organization 2020b). Our study identified that there is a significant gap in the knowledge among the general population about COVID-19. There is a need to improve individuals’ knowledge about COVID-19 preventive measures through continuous health education.

In our study, around 22.1% (n= 1,161) of the participants reported that they have used or are currently using herbal products or nutritional supplements during the COVID-19 pandemic to protect themselves from the disease. Herbal products and food supplements are used worldwide either to enhance health or to help with health-associated problems. The shortage of new and e□cient pharmacological treatments to combat viral and drug-resistant infections has turned attention to medicinal plants as a potential source of effective treatment for disease. The rapid spread of the recently emerged COVID-19 pandemic, together with the absence of an effective vaccination or treatment for COVID-19, has elevated the fear, panic, and anger among the general public (Liu et al. 2020; World Health Organization 2020a). As might be expected, individuals exposed to the dangers of such infections look for self-care measures and turn to unconventional remedies such as herbal products and food supplements to reduce the risk and make the situation more controllable (Saudi Ministry of Health 2020; Saudi Center for Disease Prevention and Control 2020; World Health Organization 2020c). The shortage of new and e□cient conventional drug treatments to combat drug-resistant infectious diseases (Zahn et al. 2019) and infections has turned attention towards medicinal plants as a potential source of novel drugs. The use of herbal products among patients has become popular, and much literature has reported its use in the case of threatening emerging infectious diseases such as SARS and MERS (Hwang et al. 2020).

Better knowledge about COVID-19 preventive measures was associated with being female, having secondary school education or higher education (PhD or a master’s degree). Unlike previous studies which showed no significant difference in knowledge about COVID-19 between males and females (Naser et al. 2020), our study found that females have better knowledge compared to males. This could be due to the difference in the assessment tool used, and the time of conducting the study as the knowledge level could increase with time during the pandemic due to continuous educational campaigns carried out by the governmental authorities. The results of previous studies were in line with ours regarding the higher level of knowledge among individuals with a higher level of education (Naser et al. 2020; Zhong et al. 2020).

Overall, knowledge about COVID-19 transmission and preventive measures among our study population was moderate, which could lead to a higher probability of risky practices during the COVID-19 pandemic. This indicates that there are gaps in the knowledge of the general population and that there is a need to improve individuals’ awareness and knowledge via extensive health education strategies.

To the best of our knowledge, this is the first study in the Middle Eastern countries that investigated knowledge about COVID-19 preventive measures (specifically handwashing techniques and social distancing), and beliefs about and the use of herbal products and food supplements among the general population during the COVID-19 pandemic. A strength of our study is that a large sample of participants was recruited during this critical period. However, there are some limitations. The study design itself, a cross-sectional survey design, limited our ability to identify causality between study variables. There are limited studies that have evaluated individuals’ knowledge about the COVID-19 pandemic and the use of herbal products and food supplements worldwide and in the Middle East specifically, which limited our ability to compare our findings with Arabic-speaking countries of a similar socioeconomic level and culture. Additionally, the use of an online survey for data collection might have excluded some vulnerable populations who could not be reached and we may have missed some of the targeted population. Finally, we were not able to estimate the response rate for our questionnaire study.

## 4. Conclusion

The findings of our study demonstrated that the general population in Saudi Arabia has a moderate level of knowledge about COVID-19 transmission and preventive measures. A considerable proportion of the population reported the use of herbal products or food supplements in order to protect themselves from the disease. We suggest that policymakers provide further educational campaigns that increase population knowledge about the disease transmission routes and preventive measures. In addition, the use of herbal products should be evidence-based to ensure patient safety.

## Data Availability

The data that support the findings of this study are available from the corresponding author upon reasonable request.

## Acknowledgments

The authors would like to express their gratitude to the ministry of education and the deanship of scientific research-Najran University – Kingdom of Saudi Arabi for their financial and technical support under code number (NU/MID/18/007).

